# Esketamine-induced post-traumatic stress disorder flashbacks during treatment-resistant depression indication: is it just a side effect?

**DOI:** 10.1101/2024.01.09.24300998

**Authors:** Maud Rothärmel, Lila Mekaoui, François Kazour, Morgane Herrero, Eva-Maria Beetz-Lobono, Aiste Lengvenyte, Jérôme Holtzmann, Philippe Raynaud, Macarena Cuenca, Samuel Bulteau, Pierre de Maricourt, Thomas Husson, Emilie Olié, Bénédicte Gohier, Anne Sauvaget, Raphaël Gaillard, Raphaëlle Richieri, David Szekely, Ludovic Samalin, Olivier Guillin, Virginie Moulier, Wissam El-Hage, Andrew Laurin, Lucie Berkovitch

## Abstract

**Background:** Posttraumatic stress disorder (PTSD) is a severe and frequent affection that is highly comorbid to major depressive disorder. Comorbid PTSD and depression are usually treatment-resistant, with a high risk of functional impairment and suicide. Esketamine nasal spray is a recent validated treatment for treatment-resistant depression (TRD), but its efficacy on comorbid TRD-PTSD remains insufficiently documented. In particular, flashbacks can occur during esketamine administration and their influence on clinical outcomes is unknown.

**Objectives:** Our main objective was to describe esketamine-induced traumatic flashbacks and their impact on clinical trajectories within a sample of patients with comorbid TRD-PTSD.

**Methods:** We retrospectively collected clinical data of patients receiving esketamine nasal spray for TRD with comorbid PTSD who experienced at least one flashback of their trauma during esketamine sessions across 11 psychiatric departments.

**Results:** Between February 2020 and March 2023, 22 adult patients with TRD met inclusion criteria. In sixteen patients (72.7%) flashbacks disappeared as the sessions progressed. In six patients (27.3%), esketamine treatment was stopped because of persistent flashbacks. When esketamine was continued, clinical response was observed both for depression and PTSD (depression response rate: 45.5% and remission rate: 22.7%; PTSD response rate: 45.5% and remission: 18.2%).

**Limitations:** The retrospective design of the study and the absence of a comparator group are the main limitations of our study.

**Conclusions:** Our results suggest that the occurrence of esketamine-induced traumatic flashbacks does not hinder clinical response. On the contrary, when managed appropriately and combined with targeted psychotherapy, it could even contribute to positive outcomes.

**Highlights:**

- Esketamine nasal spray is recently validated for treatment-resistant depression
- Its efficacy on comorbid post-traumatic stress disorder is poorly documented.
- Traumatic flashbacks can occur during esketamine administration.
- Esketamine-induced traumatic flashbacks does not hinder its clinical response.

**Clinical impact statement:** Esketamine nasal spray is recently validated for treatment-resistant depression (TRD). Its efficacy on comorbid post-traumatic stress disorder (PTSD) is poorly documented. In this study, we reported the data of 22 adult patients who received esketamine nasal spray for TRD with comorbid PTSD and experienced flashbacks during esketamine sessions. These flashbacks did not appear to be a contreindication to the administration of esketamine and clinical response was observed both for depression and PTSD. Our results suggest that esketamine could be safely administered to patients with comorbid PTSD and TRD and that esketamine could lead to a substantial improvement in this population.

## 1. Introduction

Posttraumatic stress disorder (PTSD) includes persistent and recurrent symptoms of unwanted intrusive memories, avoidance behavior, changes in physical and emotional reactions, and negative changes in thinking and mood. Among intrusive symptoms, patients may experience flashbacks of the traumatic event where they feel as if it were happening again (American Psychiatric Association, 2013). PTSD lifetime prevalence is high ∼8.3% (Kilpatrick et al., 2013). Its risk increases with trauma exposure severity, cumulative traumas, particularly for interpersonal traumas, i.e., physical and sexual assault in the context of relationship (Kessler et al., 2017). PTSD is often comorbid with other mental disorders. In around half of the cases, it is associated with major depressive disorder, which is a high-risk factor for treatment-resistance (Hegel et al., 2005; Kaplan & Klinetob, 2000; Kessler et al., 1995). Additionally, comorbid depression and PTSD induce greater functional impairment (Post et al., 2011) and a three times higher likelihood of suicidality than for depression alone (Karatzias et al., 2019).

Treatment strategies for PTSD include early interventions for prevention (Kearns et al., 2012), trauma-focused therapies (Bisson et al., 2013; Lenz et al., 2017; Watkins et al., 2018), and pharmacological treatments (e.g., antidepressants and α1 blocker) (Burback et al., 2023). Trauma-centered exposure psychotherapy is the first-line recommended therapy and has shown a significant efficacy in treating PTSD (Cusack et al., 2016; Watts et al., 2013). It consists in reactivating the traumatic memory in a safe environment and to learn how to prevent anxious response in order to reconsolidate the memory without its fear component, a phenomenon called fear extinction (Foa et al., 2008). However, up to 50% of patients may not be fully responsive to a correctly conducted psychotherapy when they have access to it (Resick et al., 2017; Steenkamp et al., 2015). By contrast, pharmacological therapy has an efficacy that appears to be modest at most (Watts et al., 2013) and in any case lower than that of psychotherapy (Merz et al., 2019). Innovative treatments for PTSD are therefore strongly needed, especially as current pharmacological and psychological therapies in combination do not appear to sufficiently improve clinical outcomes (Hetrick et al., 2010)

Ketamine is a noncompetitive *N*-methyl-D-aspartate glutamate receptor antagonist whose antidepressant properties at the dose of 0.5 mg/kg have been evidenced about 20 years ago (Berman et al., 2000) and have since been robustly reproduced (Hashimoto, 2019). Ketamine has also shown promising results in comorbid PTSD with treatment-resistant depression (TRD) (Albott et al., 2018) and in chronic PTSD alone (Duek et al., 2021; Feder et al., 2014, 2021; Pradhan et al., 2017), although some replications failed (Abdallah et al., 2022). More recently, S-enantiomer of ketamine (esketamine), which was validated for the treatment of TRD – but not PTSD –(European Medicines Agency, 2021; U.S. Food and Drug Administration, 2020), was found to improve both PTSD and depressive symptoms in a cohort of patients suffering from comorbid PTSD and TRD (Rothärmel et al., 2022). In this sample, it empirically appeared that patients could experience intense flashbacks during esketamine sessions, usually followed by a strong decrease or even a resolution of their PTSD symptoms.

In this present study, we extend those findings by compiling clinical data of patients who received esketamine nasal spray for TRD with comorbid PTSD and experienced flashbacks during esketamine sessions. Our first objective was to describe esketamine-induced traumatic flashbacks and their potential influence on clinical trajectories. A secondary aim was to provide guidelines to help patients and caregivers to manage these esketamine-induced flashbacks.

## 2. Material and Methods

### 2.1. General design and participants

This study was an observational, retrospective and multicentric study comprising patients (21– 62 years) with TRD and comorbid PTSD who have received esketamine nasal spray between February 2020 and March 2023. All the patients were treated in compliance with the guidelines provided by the French regulatory authority (ANSM, *Agence Nationale de Sécurité du Médicament et des produits de santé*) and the good clinical practice for TRD patients. Several mental health services from different university hospitals were involved in the recruitment.

The inclusion criteria comprised: *i)* Age≥18 years old; *ii)* Diagnosis of moderate-to-severe TRD, in accordance with the DSM-5 criteria (Tolentino & Schmidt, 2018) for a single or recurrent episode of major depression within unipolar or bipolar disorder; *iii)* Absence of remission after an adequate dose and duration of two or more different antidepressants given during the current episode in case of unipolar disorder; lack of remission on lithium at an adequate plasmatic level (0.8 mEq/L) combined with lamotrigine (50-200 mg per day) or full-dose quetiapine monotherapy (300-600 mg/day) in case of bipolar disorder (Pacchiarotti et al., 2009); *iv)* Comorbid PTSD according to the DSM-5; *v)* Treatment by esketamine with at least one traumatic flashback during esketamine sessions.

The study was conducted in accordance with the Declaration of Helsinki. This protocol has been approved by our local ethics committee (E2022-39, CHU Rouen, July 29^th^, 2022).

### 2.2. Study procedures

All participants received esketamine nasal spray in a full or partial hospitalization setting. All patients had baseline blood tests and an electrocardiogram to determine medical stability before the initiation of esketamine. Post-administration monitoring was undertaken by a healthcare professional for at least 120 minutes. Esketamine was initiated at 56 mg and adjusted on an individual basis during the treatment period in accordance with the following treatment guidelines: weeks 1–4: 56 or 84 mg twice weekly; weeks 5–8: 56 or 84 mg once weekly; and week 9 and onwards: 56 or 84 mg once every 1 or 2 weeks (European Medicines Agency, 2021). However, psychiatrists were free to adjust treatment dosage and frequency on an individual basis, depending on the patient’s response and tolerance to the treatment.

Anamnestic data were retrospectively collected. They included sociodemographic information, history of depressive disease, treatment history for the current major depressive episode (MDE), comorbidities, level of resistance (Maudsley staging model) and current psychotherapy received by the patient during esketamine treatment (CBT for cognitive behavioral therapy, or EMDR for eye movement desensitization and reprocessing).

Details on life-traumatic events, esketamine-induced trauma flashbacks and adverse events during esketamine sessions were also collected. Psychometric assessments included Montgomery-Åsberg Depression Rating Scale (MADRS) (Montgomery & Asberg, 1979), Clinical Global Impression-Suicide Scale (CGI-SS) (Lindenmayer et al., 2003), and PTSD Checklist for DSM-5 (PCL-5) (Ashbaugh et al., 2016). They were obtained from patients’ medical records at the time of esketamine-induced flashbacks. For depression, response to treatment was defined as an improvement of more than 50% compared to baseline MADRS scores (Frank et al., 1991) and remission was defined as the resolution of depressive symptoms, no longer allowing a diagnosis of depression according to the criteria established by the DSM-5, with a MADRS score of less than 10 (McIntyre et al., 2005). For PTSD symptoms, no psychometric assessments were routinely used. We report PCL-5 whenever available and response and remission as assessed by the patient’s clinician.

### 2.3. Statistics

Statistical analyses were conducted using SPSS, version 28 (IBM, Armonk, NY). All tests were two-tailed, with a statistical significance level set at *p*<0.05. Continuous variables are expressed as mean ± standard deviation (SD), while categorical variables are reported as percentages. Fisher’s exact test was used to assess the statistical relationship between two categorical outcomes. For continuous outcomes (number of adverse effects and depression score), Mann-Whitney non-parametric test was used.

## 3. Results

### 3.1. Sample characteristics

Eleven clinical centers took part in the study. We included a total of 22 patients (72.7% with unipolar and 27.3% with bipolar disorder) who had experienced at least one flashback of their trauma during an esketamine session. The sociodemographic and clinical characteristics are reported in **Table 1**.

**Table 1.**
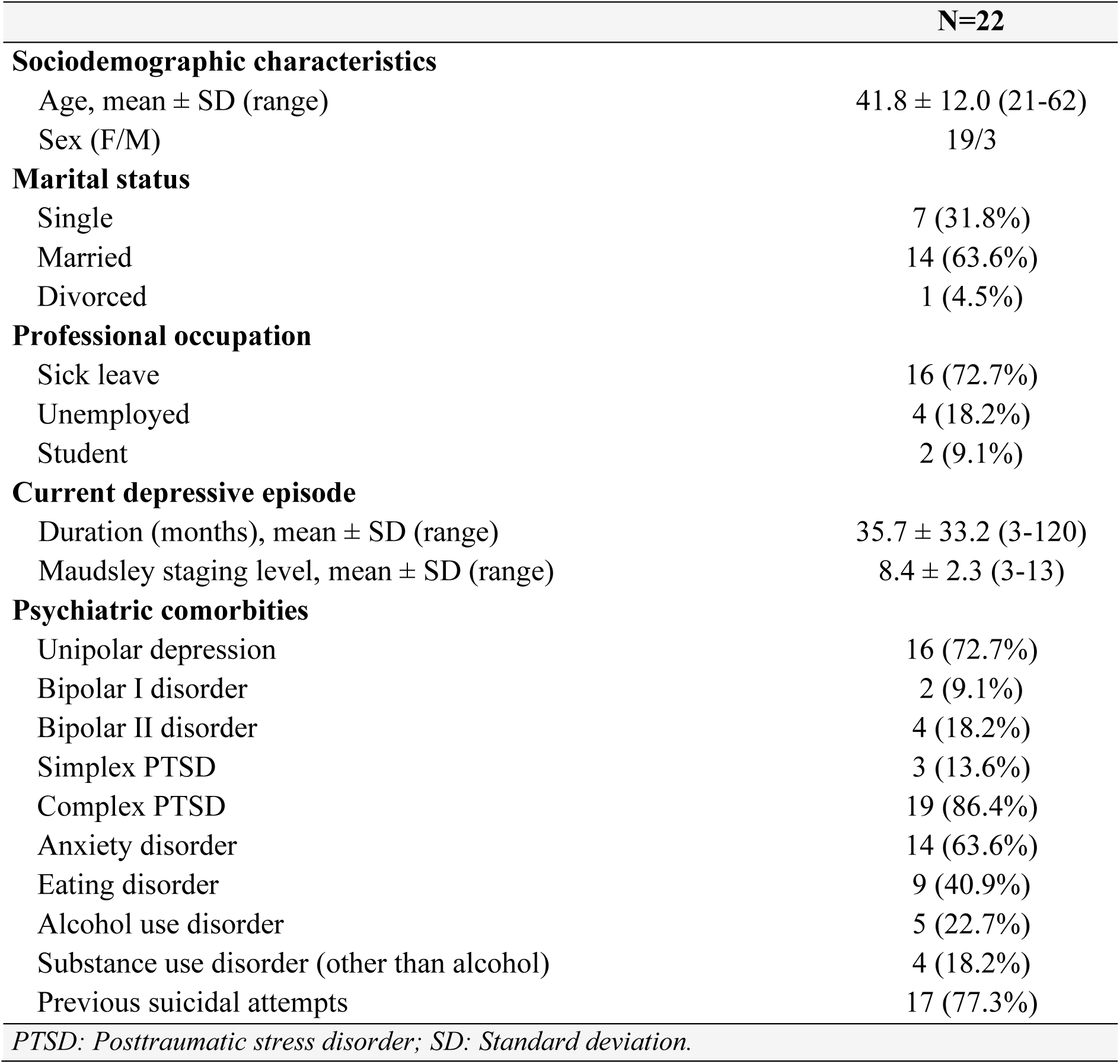
Characteristics of the sample.

Trauma causing PTSD was reported by patients as being related to sexual abuse in adulthood (n=10), sexual abuse in childhood (n=9), physical violence (n=7), discovery of a person who died (n=4), emotional negligence during childhood (n=1), forced abortion (n=1), children death (n=1) and workplace bullying (n=1).

### 3.2. Esketamine-induced traumatic flashbacks

First-induced flashbacks occurred on average during the 4.4^th^ session (SD 5.1, median 2.5, range 1-22): 40.9% of patients had flashbacks during the first session and 77.3% had flashbacks during the first five sessions.

During the week of flashbacks’ occurrence, the clinical symptoms were still high for depression (mean MADRS score 29.6 ± 9.8; n=22; range 6-50) and for PTSD (mean PCL-5 score 52.9 ± 15.0; n=9; range 31-73). The mean CGI-SS score was 2.2 ±1.6 (n=20; range 0-5).

The dosage of esketamine during the first session in which the flashbacks occurred was 56 mg in 40.9% of the patients (n=9) and 84 mg in 59.1% of the patients (n=13). Esketamine was administered during an individual session for 81.8% of the patients (n=18) and during a group session (large room with beds separated by screens) for 18.2% of the patients (n=4).

Concomitant medication prescriptions included benzodiazepines (77.3%, n=17) and antipsychotics (54.5%, n=12). All patients except one were taking antidepressants: venlafaxine (n=5), fluoxetine (n=3), duloxetine (n=3), sertraline (n=3), mirtazapine (n=3), clomipramine (n=2), paroxetine (n=2), amitriptyline (n=1) or vortioxetine (n=1). Seven patients (31.8%) were treated by lithium. Regarding psychotherapy, twelve patients (54.5%) were undergoing cognitive-behavioral therapy (CBT) or eye movement desensitization and reprocessing (EMDR).

The other adverse effects induced by esketamine were: depersonalization/derealization (without flashbacks) (59.1%, n=13), nausea and vomiting (27.3%, n=6), sensory hypersensitivity (27.3%, n=6), headache (22.7%, n=5), sedation (13.6%, n=3) and high blood pressure (4.6%, n=1).

### 3.3. Flashbacks description

The patients experienced flashbacks about the following traumatic events: rape (n=13), physical violence (n=7), discovery of the dead person (n=4), delivery with pregnancy denial (n=1) and forced abortion (n=1). In 36.4% of the patients (n=8), the flashbacks revealed the trauma to the medical team. Less than one third of patients (27.3%, n=6) had flashbacks about events they had forgotten. The flashbacks induced strong anxiety (81.8%, n=18), agitation (31.8%, n=7), or catharsis with crying and feeling of release (54.5%, n=12). According to the patients, the emotional impact was mild (4.5%, n=1), moderate (50.0%, n=11) or severe (45.5%, n=10). The impact was not associated with co-prescribed antipsychotics (Fisher’s exact test; *p*=1.00), depression score (U=58; *p*=0.895), or the number of adverse effects (U=40; *p*=0.156).

In some patients with multiple traumas, we observed that esketamine-induced flashbacks would preferentially start with the most recent trauma and progressively involve older traumas. Interestingly, one patient experienced a flashback where the traumatic scene had a different ending that was much more soothing for her than her usual flashbacks.

#### Verbatim of patients and caregivers

Patients described very vivid and distressing esketamine-induced flashbacks: “*I didn’t expect to relive the events so realistically. I could not get away from the images, I was frightened, I felt like I was present in my memories, as if my eyes were back in the body of the little girl that I was at the time… I could see the images with great precision in terms of colors, light…*”

The reaction of caregivers was considered by patients as essential: “*The support and kindness of the nurse were precious to me. The climate of trust she created allowed me to verbalize what I was experiencing.*”

Caregivers also reported an intense experience: “*She relived traumatic scenes from the past as if they were happening in the present. I tried to put her back in the present, in the office, by pointing out the place, the time and my presence with a reassuring voice. I asked her to verbalize what she was seeing and to try to change the course of the scenario by finding a way out. Once the peak passed, the patient’s face relaxed and she reported a decrease in the flow of thoughts and images.”*

### 3.4. Clinical course

Eighteen patients had finished their esketamine cure when data was collected. For those patients, the average total number of esketamine sessions was 23.2 (SD 15.6, range 4-54) and flashbacks occurred in 50.6% (SD 36.3) of the esketamine sessions.

In 72.7% of the full sample of patients (n=16), flashbacks disappeared as the sessions progressed. But, in six patients (27.3%), esketamine treatment was stopped because of persistent flashbacks on average after 6 ± 3 sessions: 3 patients had a partial response to depression but not to PTSD; 3 patients had no response to depression and PTSD. Three patients wanted to continue working on their traumas in CBT or EMDR after stopping the esketamine sessions.

As regards the clinical outcome, response and remission rates for depression were 45.5% (n=10) and 22.7% (n=5) respectively and response and remission rates for PTSD were 45.5% (n=10) and 18.2% (n=4) respectively, about 6 months after the start of treatment. A significant association was found between responses for depression and for PTSD (Fisher’s exact test; *p*=0.002) and between remission for depression and for PTSD (Fisher’s exact test; *p*<0.001).

Co-treatment by CBT or EMDR was neither associated with depression response (Fisher’s exact test; *p*=1.00) nor with PTSD response (Fisher’s exact test; *p*=0.675). Flashbacks impact was significantly associated with PTSD response (Fisher’s exact test; *p*=0.006): the responder rate was 91.7% in patients with a light to mild flashbacks impact (n=12) and only 30% in patients with severe impact (n=10).

Patients verbally reported a feeling of catharsis following esketamine-induced flashbacks in 12 cases but this feeling was not associated with PTSD response (Fisher’s exact test; *p*=0.378).

## 4. Discussion

In this retrospective observational study, we report 22 cases of patients with comorbid TRD and PTSD who received esketamine as an antidepressant treatment and experienced trauma flashbacks during at least one esketamine session. Trauma flashbacks are part of PTSD dissociative symptomatology. They can occur spontaneously and significantly contribute to patients’ suffering. At first sight, bringing those symptoms back during esketamine session could thus be considered as an adverse effect that should be avoided or impose esketamine cessation. In this cohort, esketamine was indeed interrupted in 27.3% of the patients because of a flashback occurrence during a session. In participants for whom esketamine was continued, the flashbacks gradually disappeared over the course of sessions and the patients exhibited an improvement on both depression and PTSD scores (45.5% of response and ∼20% of remission on average) without any severe adverse event. Moreover, the outcomes for depression and PTSD were significantly correlated, suggesting that esketamine could be efficient either on one of these two dimensions with beneficial consequences on the other, or simultaneously on the two dimensions.

Esketamine could contribute to PTSD healing at different levels. First, it seems to help patients to verbalize their trauma. Indeed, 36.4% of them had not previously talked about their trauma to the medical team. More importantly, esketamine may facilitate access to buried memories since 27.3% of patients reported that they had forgotten the events that reappeared during the session. Thanks to its antidepressant properties, esketamine could decrease the negative effects associated with flashbacks and act as an analgesic molecule favoring desensitization. Finally, derealization occurring during dissociation sometimes places the patient in the position of being a spectator to traumatic reviviscences, which can help to gain emotional distance.

Interestingly, in this case series, the impact of flashbacks seemed to play a role in the subsequent outcome, with a higher PTSD response rate if the impact was light or mild and a lower rate of response when the impact was severe. The profile of these two categories of patients may differ and confounding factors could have contributed both to the intensity of flashbacks impact and to the response rates, independently of any esketamine administration. Nevertheless, this result is in line with the general principles of exposure-based behavioral therapies in PTSD (Rothbaum & Schwartz, 2002): sufficiently long exposure in a safe environment allows for habituation and extinction of fear, while the persistence of anxiety at the end of the session may further sensitize patients. In this regard, we explored whether the co-prescription of antipsychotics could decrease flashbacks impact but did not observe any significant effects. Further studies are needed to assess whether co-prescriptions could help to decrease anxiety, thereby promoting extinction.

Both the access to forgotten trauma during esketamine sessions and the anti-chronologically order in which flashbacks often occur suggest that esketamine may favor memory reactivation, since relatively more recent memories are more prone to destabilization than older ones (Frankland et al., 2006). Finally, flashbacks were sometimes very intense and had a cathartic effect even if this aspect did not seem to be associated with PTSD resolution.

Because of the progressive decrease in flashbacks observed over the course of the sessions in this sample, we suggest that trauma-focused psychotherapy, with exposure-extinction procedure, could be delivered after esketamine administration to potentiate esketamine effect. This proposal is also supported by previously documented neurocognitive properties of ketamine, notably its effects on emotional processing and on synaptic plasticity.

Indeed, first, ketamine has been shown to decrease amygdala reactivity to negative emotional stimuli in healthy controls (Abel et al., 2003; Scheidegger et al., 2016) and in patients with major depressive disorder (C.-T. Li et al., 2016). In patients with PTSD, ketamine’s beneficial effects are correlated with an increased top-down inhibition of the amygdala by the ventromedial prefrontal cortex during emotional face-viewing, and ketamine efficacy is higher in patients with low prefrontal inhibition of amygdala responses to faces at baseline (Norbury et al., 2021). In this view, esketamine would normalize the increased sensitivity to negative emotional stimuli in depression and PTSD (Groenewold et al., 2013; Hayes et al., 2012) by restoring the prefrontal control over the amygdala. Psychotherapeutic techniques could even be proposed during esketamine sessions to better manage the advent of flashbacks and decrease fear memory reconsolidation. For instance, it has been suggested that the link between dissociation and mindfulness may be mediated by attention and emotional acceptance (Vancappel et al., 2023), and that specific exercises targeting both processes would be efficient to treat dissociative symptoms (Bishop et al., 2004). In order to foster acceptance, caregivers can use the “train metaphor” inspired from EMDR techniques (Shapiro, 1989). The patient is invited to consider that he is like the passenger on the train watching the landscape passing by. The landscape can be composed of emotions, sensations, thoughts, and images. The patient is encouraged letting this landscape pass. Then, the patient is invited to pay attention to the present moment by noting what she/he currently perceives with his five senses and from her/his internal experience (emotions, sensations, thoughts). This “dropping anchor” technique (Harris, 2017) derived from Acceptance and Commitment Therapy (ACT) (Hayes et al., 2012), allows the patient to be here and now, intentionally, without filter or judgment.

Second, under esketamine, flashbacks occur when patients are in a different state of mind and their subjective feelings and memories may be more easily reshaped. Importantly, its effect on memory seems to be highly dependent on the delay with encoding. Indeed, several studies found that ketamine could inflate memory retention or impede fear extinction when administered before or right after fear conditioning in experimental psychology paradigms (Corlett et al., 2013; Honsberger et al., 2015; Morena et al., 2017), with potential detrimental effect on PTSD outcomes (Juven-Wetzler et al., 2014; Schönenberg et al., 2005, 2008). On the contrary, we posit that ketamine elicits ancient memories destabilization through subjective experience of dissociation. Indeed, at a neurobiological level, ketamine acts as an NMDA antagonist, resulting in a secondary glutamate release and *in fine* in brain excitation and an increased synaptic plasticity (Duman et al., 2016), notably in the prefrontal cortex (N. Li et al., 2010; Pham & Gardier, 2019). Synaptic plasticity and glutamate receptor modulators have been suggested to play a role in memory destabilization and reconsolidation (Finnie & Nader, 2012; Monfils et al., 2009). More specifically, ketamine seems to facilitate fear memory extinction (Duek et al., 2021; Philippens et al., 2021; Wei et al., 2020) possibly through a decrease of fear memory reconsolidation (Duclot et al., 2016) or a decreased access to contextual details during memory retrieval (Honey et al., 2005).

In line with previous proposals (Fattore et al., 2018), we therefore suggest that esketamine constitutes a pharmacological tool that could foster trauma-resolution through different mechanisms: 1) destabilizing memory and increasing the advent of flashbacks, 2) enhancing patients’ control over their negative emotions during flashbacks, a mechanism that could be reinforced through dedicated psychotherapeutic interventions and 3) reducing fear memory reconsolidation during reactivation and thereby favoring extinction. In this perspective, we strongly recommend that PTSD should not be considered as an exclusion criterion for esketamine treatment and that patients with PTSD receiving esketamine should be informed in advance of the possible advent of flashbacks during the sessions. Then, they could be proposed a psychotherapeutic support during administration to maximize benefits. Caregivers must also be prepared for this type of reaction.

Interestingly, another molecule with a different pharmacological mechanism, 3,4-methylenedioxy-methamphetamine (MDMA), significantly reduces PTSD and depressive symptoms when associated with psychotherapy (Mitchell et al., 2021; Smith et al., 2022). MDMA has specific cognitive effects that are particularly relevant to alleviating PTSD symptoms. For instance, it enhances emotional empathy (Hysek et al., 2014; Kuypers et al., 2017), increases self-compassion (Kamboj et al., 2015) and has several pro-social effects (Bedi et al., 2009; Hysek et al., 2014) that could facilitate the therapeutic alliance and psychotherapeutic work. Like ketamine, it decreases amygdala reactivity to negative emotional stimuli (Bedi et al., 2009; Gamma et al., 2000) and promotes fear memory extinction (Hake et al., 2019; Young et al., 2015), suggesting that those changes play a crucial role in treatment efficacy. The interest of reactivating memories and to modulate them with other therapeutic interventions has been investigated with mixed results, e.g., β-adrenergic receptor antagonist propranolol (Brunet et al., 2018; Raut et al., 2022; Vaiva et al., 2003), transmagnetic stimulation (Isserles et al, 2013, but: Isserles et al., 2021) and electroconvulsivotherapy (Gahr et al., 2014; Kroes et al., 2014, but: Tang et al., 2021).

On another note, ketamine has already been proposed as a “maximizer” of cognitive behavioral therapy in other psychiatric affections, such as obsessive-compulsive disorder (Rodriguez et al., 2016), depression (Bottemanne et al., 2021; Dore et al., 2019; Hasler, 2020) and addictions (Grabski et al., 2022; Krupitsky et al., 2002), notably through its ability to rewrite maladaptive memories (Das et al., 2019). Our results suggest that it could also be the case for PTSD.

Despite the perspectives that our study provides, several limitations should be mentioned. First, it was not designed to compare the outcomes for patients with comorbid TRD and PTSD based on whether or not they would experience traumatic flashbacks under esketamine; or to assess the prevalence of esketamine-induced trauma flashbacks among this population. Indeed, data were retrospectively collected after the empirical observation that some patients had such flashbacks during an esketamine administration and felt subsequently relieved. Instead, our study intends to highlight the existence of this phenomenon and emphasizes that these patients may have positive outcomes if esketamine is not interrupted. Further studies with a systematic and prospective data collection are required to evaluate the prevalence of esketamine-induced trauma flashbacks and its potential role as a predicting factor of response. Second, our study does not allow to compare esketamine efficacy in comorbid TRD and PTSD and TRD alone. The response rate looks quite similar to that of a real-world study that assessed the efficacy of esketamine in TRD in a French cohort (45.5% in our study versus 47.8% in this study on TRD) (Samalin et al., 2022). By contrast, the rate of remission appears to be lower (22.7% in our study versus 37% in the TRD study), possibly because comorbid PTSD leads to the persistence of residual symptoms that are more intractable. Larger cohorts including both patients with TRD alone and comorbid PTSD and TRD could in the future perform those comparisons.

## Conclusion

In summary, this case series shows that esketamine could be safely administered to patients with comorbid PTSD and TRD even if flashbacks occur during the sessions. Comorbid TRD and PTSD usually have a poor prognosis (Karatzias et al., 2019; Panagioti et al., 2012; Post et al., 2011) but our study suggests that esketamine could lead to a substantial improvement in this population (see also: Rothärmel et al., 2022). Indeed, thanks to its specific neurocognitive effects, esketamine may favor the advent of flashbacks but also facilitate trauma-resolution by decreasing negative emotions associated to them and promoting fear extinction, in particular if psychotherapy is simultaneously delivered.

## Data Availability

All data produced in the present study are available upon reasonable request to the authors

## Acknowledgments

We thank all the participants. We also thank Gérald Adler, Cherifa Benosman, Julien Corniquet, Wendy Chevallier, Graziella Guesdon, Carine Kerninon, Corinne Lagniez and and Julie Mougeolle for assistance in esketamine administration.

LB thanks the *Fondation Bettencourt-Schueller, the Philippe Foundation*, the *Foundation L’Oréal-Unesco* and the National Institute of Mental Health (R01MH116038 and U01MH121766) for their support.

## Author contributions statements

*Conceptualization* : MR, LM, LB ; *Formal analysis* : VM ; *Investigation* : MR, LM, ALa, FK, MH, E-VB-L, Ale, JH, PR, MC, TH, RR, ; *Methodology* : MR, LM, LB, AS ; *Project administration* : MR, LM ; *Supervision* : OG, AS, RG, WE, SB, PM, EO, BG, DZ, LS ; *writing original draft* : MR, LB ; *Writing—review & editing* : ALa, SB, AS, RG, WE, TH, LS.

